# Functional response to a microbial synbiotic in the gastrointestinal system of constipated children

**DOI:** 10.1101/2022.04.07.22273329

**Authors:** Braden T. Tierney, James Versalovic, Alessio Fasano, Joseph F Petrosino, Bruno Chumpitazi, Emeran A. Mayer, Jared Boetes, Gerard Smits, Shanthi G. Parkar, Noah Voreades, Ece Kartal, Peter A. Bron, Gregor Reid, Raja Dhir, Christopher E. Mason

## Abstract

**Background:** Oral microbial therapy has been studied as an intervention for a range of gastrointestinal and immunological disorders. Though emerging research suggests microbial exposure may intimately affect the gastrointestinal system, motility, and host immunity in a pediatric population, data has been inconsistent and variable, with the majority of prior studies conducted in neither a randomized nor placebo-controlled setting. The aim of this placebo-controlled study was to evaluate efficacy of a synbiotic (a prebiotic and rationally-defined microbial consortia) on increasing weekly bowel movement frequency in constipated children.

**Methods:** Sixty-four children (3-17 years of age) were randomized to receive a synbiotic composition (n=33) comprised of mixed-chain length, prebiotic oligosaccharides and nine microbial strains or placebo (n=31) for 84 days. Stool microbiota was analyzed using shotgun metagenomic sequencing on samples collected at baseline (T1) and completion (T2). The primary outcome was change from baseline of Weekly Bowel Movements (WBMs) in children compared to placebo.

**Results:** Treatment with a multi-strain synbiotic significantly (p < 0.05) increased the number of WBMs in children with low bowel movement frequency (< 4 WBMs and < 5 WBMs), irrespective of broadly distinctive microbiome signatures at baseline. Metagenomic shotgun sequencing revealed that low baseline microbial richness in the treatment group significantly anticipated improvements in constipation (p = 0.00074).

**Conclusions:** These findings suggest the potential for (i) multi-species synbiotic interventions to improve digestive health in a pediatric population and (ii) bioinformatics-based methods to predict response to microbial interventions in children.

**Impact:** Synbiotic microbial treatment exerted functional improvements in the number of spontaneous Weekly Bowel Movements in children compared to placebo

Intervention induced a significant bifidogenic effect in children compared to placebo

All administered probiotic species were enriched in the gut microbiome of the intervention group compared to placebo

Baseline microbial richness demonstrated potential as a predictive biomarker for response to intervention

## Background

Recent advances in microbiome tools (e.g. culturing, bioinformatics) have enabled a deeper understanding of microbial ecology and the gut microbiome’s role in human health and wellbeing. Gastrointestinal microbes exert functional influence on the host through a range of metabolic and immunological mechanisms and the host shapes resident microbial communities through diet, nutrition, lifestyle, and medication.^1–3^ The composition of the human gut microbiome has been identified as playing a role in regulating bowel movements in children, including functional constipation (FC), which is characterized by infrequent bowel movements and associated phenotypes therein (e.g. stool consistency, pain when defecating, bloating).^4–9^ FC afflicts about 25% of children visiting pediatric gastroenterologist practices in the United States.^10^ Symptoms of pediatric FC frequently persist into adolescence and adulthood despite treatment with laxatives, indicating the need for alternative treatment paradigms.^11^

Gut microbiota are suggested to influence bowel movement frequency through multiple mechanisms. These include ligand-receptor type interactions with competitive exclusion of pathogens, generation of antibacterial substances, setting an anti-inflammatory tone to the gut environment, signaling effects that influence the enteric nervous system, and breakdown of fiber to generate short chain fatty acids (SCFAs) that improve gut function.^12–15^ Given the reported link between microbial metabolism and FC, modulation of the gut microbiota in children may lead to beneficial clinical outcomes for those experiencing bowel distress.

A number of pilot studies have been conducted to identify if microbial therapies can improve the quantity and quality of Weekly Bowel Movements (WBMs). These have included supplementation of live bacteria (i.e. candidate probiotics), ingredients to support the growth of beneficial organisms (i.e. prebiotics), or a combination of the two (i.e. synbiotics). In children, an intake of inulin-type fructans has been associated with softer stool consistency^27^ (a component of functional constipation) as well as protection against gastrointestinal infection,^28^ antibiotic-induced bifidobacterial depletion,^29^ weight gain,^30^ and mineral malabsorption.^31^ At least some of these benefits are attributed to fructan-mediated bifidogenic effect.^28,32^ The by-products of bifidobacterial metabolism, predominantly organic acids, are further known to cross-feed secondary feeders, mainly butyrogenic bacteria from *Lachnospiraceae* and *Ruminococcaceae*.^*33,34*^ The increase in a more diverse microbiome helps to build a greater heterofermentative capacity, thus nurturing a more beneficial microbial consortium.

For specific strains, there have been statistically significant, beneficial outcomes in human trials.^16,17^ Specific genera of interest empirically include lactobacilli and bifidobacteria.^18,19^ For prebiotics, fiber -- which is a key microbial nutritional substrate -- is potentially effective in improving bowel movement frequency, especially in children.^20–22^ That said, meta-analyses evaluating the relationship between nutritional inputs and constipation relief showed substantial heterogeneity in results.^19,23,24^ However, these studies have predominantly been in non-pediatric cohorts, they have lacked a placebo arm, and they tend to use single-organism interventions.^25,26^

Consequently, there is a need to test in placebo-controlled trials the efficacy of microbial therapies on reducing pediatric constipation and its associated symptoms. Here, we do so in a pilot study to determine the impact of a nine-strain (eight species) synbiotic formulation (with the prebiotic comprising of mixed-chain length oligosaccharides) on ameliorating constipation.

## Methods

### Study design and primary objective

The clinical trial was IRB-approved, randomized, double-blind and placebo-controlled with two parallel arms. Following a run-in period of 14 days, subjects were randomly assigned into an intervention and placebo arm for a duration of 84 days. Constipated was defined as having fewer than 4 WBMs, whereas low WBMs was defined as having fewer than 5. The primary objective of the study was to assess the change from baseline to Day 84 in weekly frequency of spontaneous bowel movements between subjects receiving placebo and those receiving a multistrain synbiotic.

### Randomization and patient selection

One hundred and twenty-one healthy male/female subjects were assessed for eligibility (Fig 1, Supp Table 1). Exclusion criteria included obesity, pregnancy, lack of parental consent or ability to collect data during the study, and a lack of medical history. Thirty subjects were excluded due to exclusionary self-reported medications or body-mass-index measurements. The remainder underwent a 14 day run-in to establish baseline weekly bowel movements as a 7 day average with daily reporting. Variation was observed in the parental reported baseline WBMs during the 14 day run-in period. The results showed a heterogenous pediatric population with highly variable baseline WBM frequency, and many subjects did not meet our definition of constipation. Thirty-eight of the 64 subjects who completed through day 84 had fewer than 5 WBMs, and 21 had less than 4 WBMs. The randomization resulted in 43 subjects in the synbiotic arm and 48 receiving placebo. Parents reported in a logbook the daily frequency and consistency of their child’s stool and any adverse events or use of medications. The subjects were asked to not consume any additional probiotic supplements or foods during the study period. WBMs were reported to the study coordinators twice weekly throughout the 84-day intervention period. While first-dosing was administered in 91 children, 27 children did not complete through day 84. These study participants or their legal guardian(s) received a termination notice upon which clinical product or placebo was returned to the study coordinator for prompt disposal. All data for these non-completing 27 children at baseline were dropped from the analysis and baseline microbiome samples destroyed.

**Table 1:** Cohort summary, response rate, and analyses of clinical outcomes.

| Sample | Endpoint | Total Patients | Response (%) |  | P-value |
| --- | --- | --- | --- | --- | --- |
|  |  |  | Intervention | Placebo |  |
| Constipated < 4 | 1 WBM increase | 21 (13 active, 8 placebo) | 62% | 25% | 0.104 |
| Constipated < 4 | 2 WBM increase | 21 (13 active, 8 placebo) | 62% | 13% | 0.0398* |
| Constipated < 4 | 3 WBM increase | 21 (13 active, 8 placebo) | 54% | 13% | 0.0612 |
| Constipated < 5 | 1 WBM increase | 38 (19 active, 19 placebo) | 63% | 37% | 0.0877 |
| Constipated < 5 | 2 WBM increase | 38 (19 active, 19 placebo) | 63% | 32% | 0.0342* |
| Constipated < 5 | 3 WBM increase | 38 (19 active, 19 placebo) | 42% | 11% | 0.0483* |
| Not Constipated ≥ 5 | 1 WBM increase | 26 (14 active, 12 placebo) | 36% | 58% | 0.1710 |
| Not Constipated ≥ 5 | 2 WBM increase | 26 (14 active, 12 placebo) | 36% | 33% | 0.8855 |
| Not Constipated ≥ 5 | 3 WBM increase | 26 (14 active, 12 placebo) | 21% | 25% | 0.6628 |
| All | 1 WBM increase | 64 (33 active, 31 placebo) | 52% | 45% | 0.7306 |
| All | 2 WBM increase | 64 (33 active, 31 placebo) | 52% | 32% | 0.1654 |
| All | 3 WBM increase | 64 (33 active, 31 placebo) | 33% | 16% | 0.1473 |

**Figure 1:**
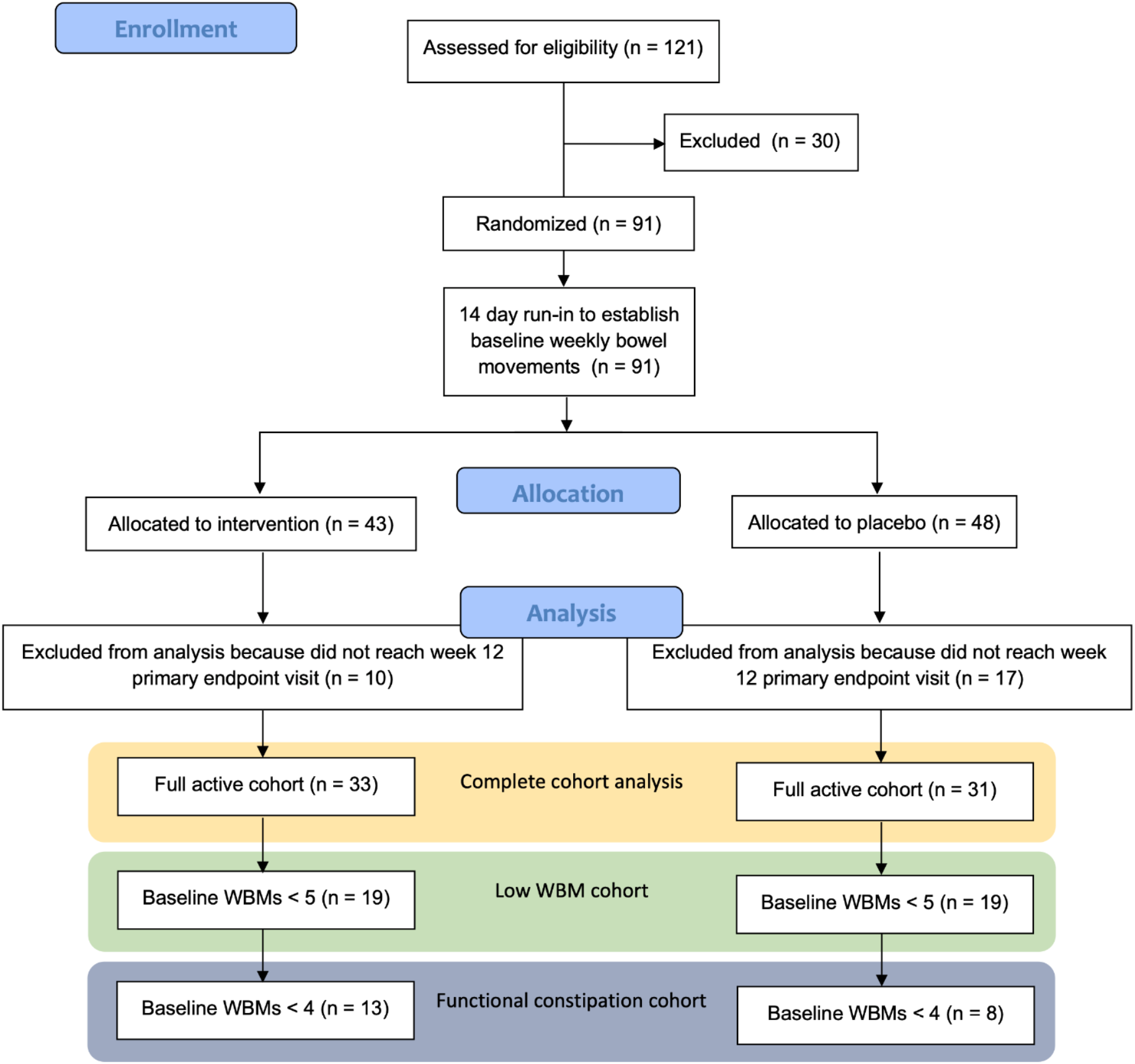
Randomized, placebo-controlled clinical design. Diagram indicating clinical trial design and execution.

### Intervention

The interventional composition consisted of 6.2 grams of mixed-chain length prebiotic substrates suspended in a single sachet with nine microbial strains (and within them, eight species): *Bifidobacterium breve* B632, *Bifidobacterium breve* BR03, *Bifidobacterium lactis* BPL1, *Bifidobacterium longum* ES1, *Lacticaseibacillus* (formerly *Lactobacillus*) *casei* BPL4, *Lacticaseibacillus* (formerly *Lactobacillus*) *rhamnosus* GG, *Lactobacillus acidophilus* NCFM, *Bifidobacterium animalis* subsp. *lactis* Bi-07, and *Ligilactobacillus* (formerly *Lactobacillus*) *salivarius* LS01. The placebo was composed of weight-, color-, and taste-matched tapioca maltodextrin and fructose. Both active and placebo were packaged in identical, unmarked sachets and shipped directly to the subject’s home to be administered daily under the supervision of the child’s legal, consenting guardian.

### Statistical analysis of clinical data

Analyses of clinical outcomes were performed using SAS^35^ software, version 9.4. Plots were produced using R^36^ version 4.1.1. Descriptive summaries included the mean, standard deviation, median, and 95% confidence interval for continuous variables, and counts, percentages, and 95% confidence intervals for categorical variables. Confidence intervals for binary endpoints were calculated using the Clopper-Pearson^37^ exact method. A two-tailed alpha of 0.05 or smaller was considered to be statistically significant.

In relation to WBM frequency, we considered two sub-cohorts as presenting with clinically relevant bowel movement patterns at baseline, defined as children with < 4 WBMs and children with < 5 WBMs. Endpoints assessed were (i) increase of ≥ 1 WBM from baseline to Day 84, (ii) increase of ≥ 2 WBM from baseline to Day 84, (iii) increase of ≥ 3 WBM from baseline to Day 84, and (iv) KINDLE quality of life (QOL questionnaire) as a standardized tool to assess adverse reactions and tolerability. We additionally measured if children with > 4 WBMs experienced a change in WBM frequency as a function of treatment.

Binary endpoints (1, 2, and 3) were analyzed by logistic regression, with adjustment for age and baseline number of WBM. The continuous endpoint 4 was analyzed with covariance (ANCOVA), with adjustment for baseline and age. Cross-tabulations of the three responder endpoints with treatment provided counts and percentages. Summary statistics were provided for change in number of WBMs by subject groupings.

### Metagenomic sequencing

Fecal samples were extracted by Diversigen with PowerSoil Pro (Qiagen) automated for high throughput on the QiaCube HT (Qiagen), using Powerbead Pro Plates (Qiagen) with 0.5mm and 0.1mm ceramic beads. Samples were quantified with Quant-iT PicoGreen dsDNA Assay (Invitrogen). Libraries were prepared with a procedure adapted from the Illumina DNA Prep kit (Illumina) and sequenced on an Illumina NovaSeq using paired-end 2×150 reads (Illumina) targeting a mean read depth of at least 4 million reads per sample.

### Diversity and microbiome feature abundance quantification

All shotgun metagenomic sequencing was quality-controlled prior to analysis. We executed all quality control with a combination of bbtools^38^ and Bowtie2^39^. We used bbmap to clump (clumpify.sh, optical=f, dupesubs=2, dedupe=t) reads and removed adapter contamination with bbduk (qout=33 trd=t hdist=1 k=27 ktrim=“r” mink=8 overwrite=true trimq=10 qtrim=‘rl’ threads=10 minlength=51 maxns=-1 minbasefrequency=0.05 ecco=f). We used repair.sh (also from bbtools, default settings), to repair any files with mismatched reads. We aligned to the human reference genome (hg38) using bowtie2 (--very-sensitive-local) to remove human sequences from stool samples. Finally, tadpole (mode=correct, ecc=t, ecco=t) was used to correct sequencing errors.

Annotation of microbial taxa, pathways, and gene family abundances was performed using MetaPhlAn3 and HUMAnN3 running the default settings.^40^ For a robustness check in our diversity and richness analysis, we used Kraken2 (default settings) as an alternative method for computing the number of Operational Taxonomic Units (OTUs) in each sample.^41^ We used Bracken to compute the abundances of each OTU at each taxonomic stratification (phyla, classes, orders, families, genera, species).^42^

Shannon and Simpson diversity were computed for each phylogenetic level with the vegan package in R. We computed taxonomic richness by summing the total number of observed taxonomic units for a given phylogenetic level (e.g. the total number of species with non-zero abundance). In accordance with the literature, we did not rarify microbiome data prior to diversity, richness, or other analyses.^43^

### Quantification and comparisons of bifidogenic and probiotic strain abundance

We took a metapangenomic approach to compute the relative abundances of particular strains of interest in our active versus treatment groups at baseline and endpoint. Specifically, these organisms were (i) microbial strains administered in active and, (ii) all members of the *Bifidobacterium* genus (NCBI taxid = 1678) with representative genomes on NCBI’s assembly database. The latter was used to assess a potential bifidogenic effect referred to as bifidogenicity or enrichment in bifidobacteria. We acquired the microbial genome sequences from both the original groups involved in strain isolation as well as from NCBI’s assembly database.

We used a portion of the Anvi’o’s metapangenomic workflow^44,45^ to compute the abundance of each microbial genome in our sequenced and quality-controlled metagenomic data. This approach consists of quantifying the abundance of each gene in a genome in a given metagenome by alignment (https://merenlab.org/data/prochlorococcus-metapangenome/). Due to this being a whole-genome-alignment-based approach and therefore possibly lacking the resolution to resolve strain-level differences simply based on mapped reads, we did not attempt to distinguish between genomes of the same species (i.e. *B. breve*). To determine a single summary statistic for each genome’s abundance, averages across the logged (adding 0.00001 to account for zero values) total abundance of each gene were computed by Anvi’o’s “anvi-script-gen-distribution-of-genes-in-a-bin” function for all genomes of interest. Specifically, the gene-by-gene abundances we used were in the GENE-COVs.txt files generated by this step.

Specifically for the *Bifidobacterium* analysis, the same approach was taken using public data from NCBI (complete genomes annotated as *Bifidobacterium*). Organisms were only selected with (i) a different species-level annotation than any of the members of the administered consortia and (ii) at least 50% of the genes in their genome represented in at least 3 samples (i.e. having non-zero abundance). This removed 81 genomes from those downloaded from NCBI. Subsequently, Anvi’o was employed to compute the abundance of each of these genomes in the dataset.

### Metagenome-Association-Study (MAS) on microbiome feature abundances

We executed a MAS between microbial feature (i.e. taxon/pathway) abundance and responder status, treatment, bloating, pain, and weekly/change in bowel movements. We looked at associations between these clinical variables and microbial feature abundances at baseline/endpoint where relevant (e.g. we did not compute the association between endpoint microbiome abundances and baseline WBMs). Prior to running the analysis, we took the natural log of all microbiome features and added a fudge factor of 0.00001. We additionally removed features that occurred in fewer than 3 samples.

For binary outcome variables, we used logistic regression adjusted for baseline WBMs and age. For all other regressions (with continuous dependent variables), we used linear regression with a gaussian link function. The only exception to adjusting strategy was when baseline weekly WBMs was the outcome variable, in which case we did not include it as an independent variable as well. For each dependent variable, we adjusted for multiple hypothesis correction using the Benjamini-Yekutieli procedure.

## Results

### Synbiotic use increases weekly bowel movements in constipated children compared to placebo

We aimed to estimate the increase in WBMs across the entire cohort, including both constipated and non-constipated individuals. We recruited 121 individuals, 30 of which were excluded prior to the study beginning (see *Methods)*. 64 (33 active and 31 placebo) returned for the Day 84 timepoint (Fig 1, Table 1, Supp Table 1). Logistic regression adjusted for age and baseline WBMs was used to measure the change in constipation in the placebo versus treatment groups. We stratified outcomes by changes in WBMs between baseline and Day 84, comparing those who (i) experienced increased weekly bowel movements (WBMs) by at least 1 relative to baseline (1 WBM), (ii) 2 relative to baseline (2 WBM), and/or (iii) 3 relative to baseline (3 WBM). Across the entire cohort, we were unable to identify significant differences between placebo and treatment participants for any of these three cutoffs, indicating that healthy individuals did not experience a further increase in WBMs.

We aimed to test if treatment improved bowel movement frequency in the 21 children with constipation as defined by < 4 WBMs. Overall, 61.5% individuals in the treatment group experienced an increase of at least 1 WBM (compared to 25% in the placebo group). We identified statistically significant, positive associations between treatment and increase in bowel movement frequency at 2 WBMs (p = 0.0398). By this metric, 8 individuals in the active improved, whereas only 1 in the placebo group did. 1 WBM and 3 WBMs were trending towards statistical significance (1 WBMs: p = 0.104; 3 WBMs p = 0.0612).

We executed one final analysis, identifying a second cut off point of < 5 WBMs (N = 38, 19 active, 19 placebo) while still including only individuals with low numbers of WBMs. Within the treatment group, 63.2% of individuals experienced an improvement of at least 1 WBM (compared to 36.8% within the placebo arm). A total of 42.1% participants in the treatment arm experienced an increase of at least 3 WBMs (compared to 10.5%, in the placebo arm). We identified positive, statistically significant associations between treatment and increases in WBMs in the 2 WBMs (p = 0.034) and 3 WBMs endpoints (p = 0.048).

### Probiotic strains are detectable in increased abundance in the treatment arm and yield a bifidogenic effect

We next aimed to investigate microbiome changes as a function of treatment and response to treatment. We received stool samples from 52 individuals at both the baseline and day 84 timepoints. We carried out shotgun sequencing on these samples in an effort to estimate their microbiome composition as a function of treatment and changes in constipation.

We queried if the 8 microbial species present in the intervention were detectable at greater abundances in the treatment versus the placebo group at trial endpoint (Fig 1). We aligned quality-controlled metagenomic sequencing reads to the Open-Reading-Frames (ORFs) in each strain’s draft genome and computed the overall average relative abundance of ORFs on a per-organism standpoint. We found no statistically significant shifts in strain abundance between baseline and endpoint in the placebo group. By contrast, we detected statistically significant (p < 0.05) shifts in species abundance for seven out of eight probiotic species in the treatment group (Fig 2) with the eighth species (*L. salivarius*) trending towards significance (p = 0.059).

**Figure 2:**
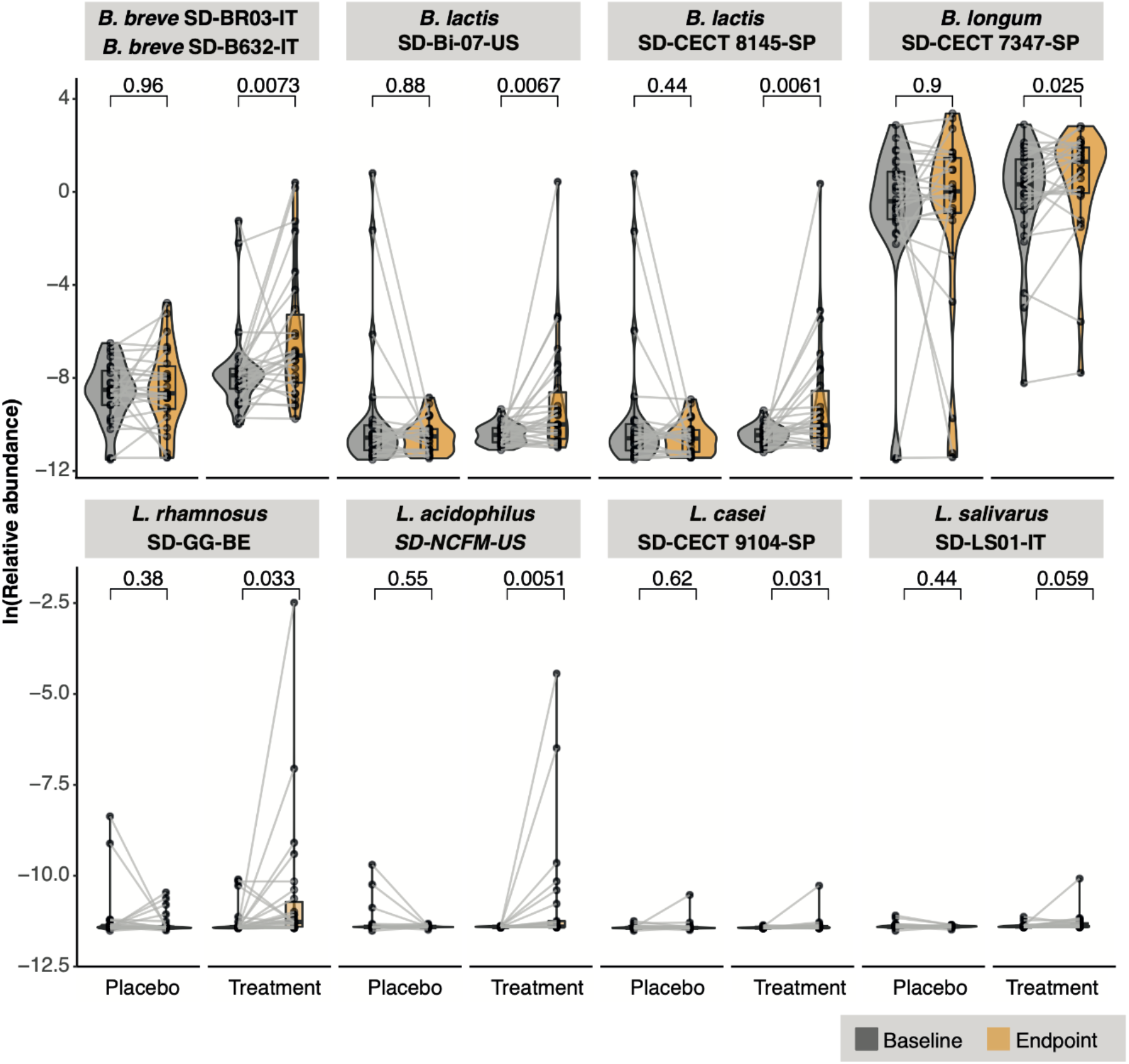
Strain relative abundance before and after treatment. Detection of strains at baseline vs. endpoint in the treatment vs placebo groups.

We performed a similar analysis to test if the administration of the synbiotic yielded a bifidogenic effect: an increase in the genus *Bifidobacterium* after treatment compared to placebo. We focused on the differential detection of strains from species other than those in the intervention consortia, selecting only for *Bifidobacterium* species where >50% of their genomes were detected in three or more samples. This yielded a total of four species of interest that were not present in the intervention (in addition to two that were). None significantly changed in overall genome abundance between the baseline and endpoint timepoints in the placebo group, whereas only one (*B. bifidum)* was approaching significance in this regard (Wilcoxon p = 0.053) in the treatment cohort (Fig 3). *Bifidobacterium* strains of the same or similar species as those in the intervention were all statistically significantly (p < 0.05) increased at the study endpoint compared to baseline in the treatment group (whereas they were not in the placebo group).

**Figure 3:**
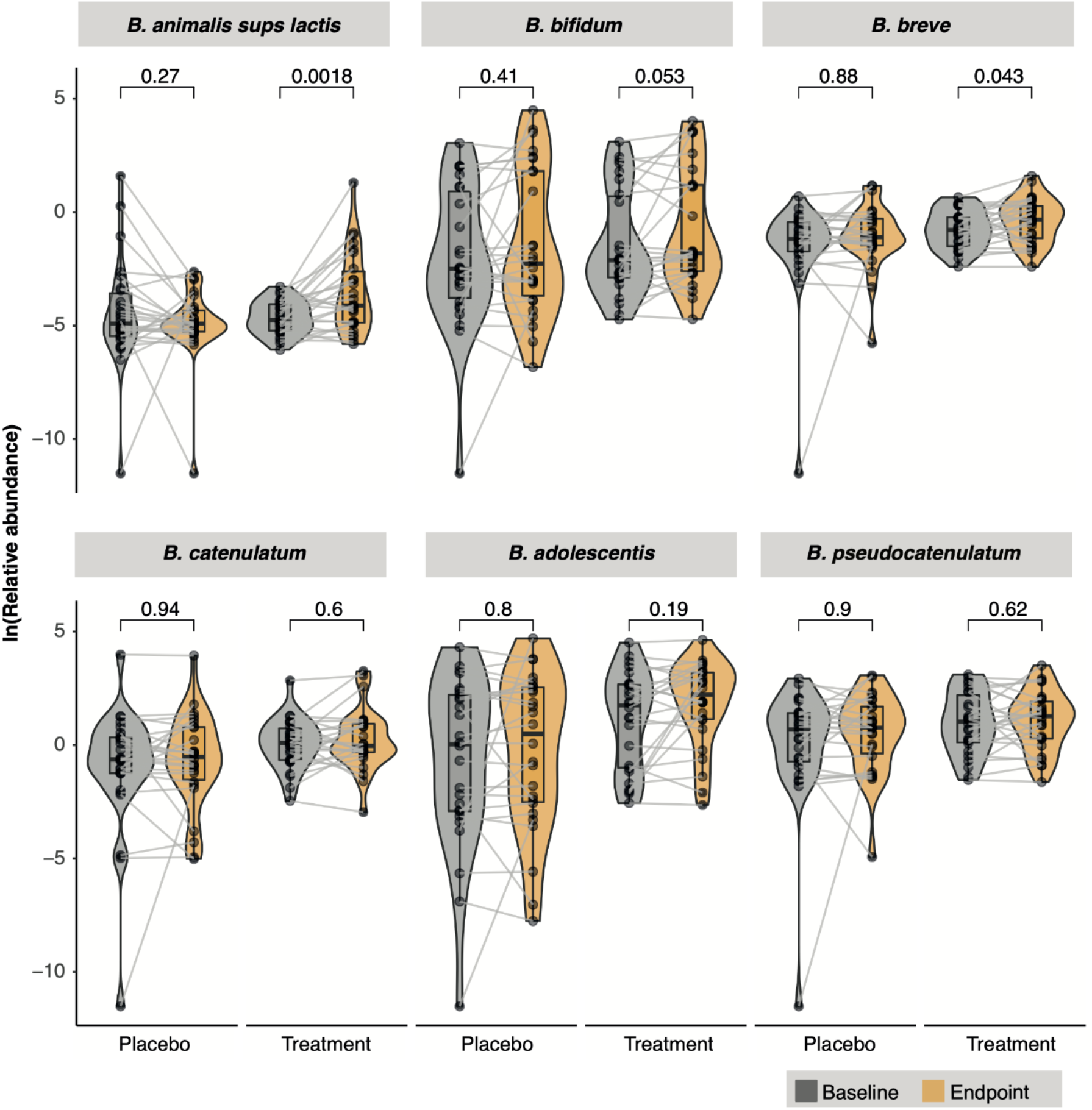
Bifidogenic effect across timepoints and study groups. Detection of members of the genus *Bifidobacterium* at baseline and endpoint in the treatment vs placebo groups.

### Limited features are significantly altered in treatment or constipation-associated phenotypes in this study

Via a discovery-driven Microbiome-Association-Study (MAS), we aimed to determine if the abundance of specific taxonomies or pathways were altered at baseline versus the endpoint as a function of treatment, bloating, pain, and weekly bowel movements. Adjusting for age and baseline WBMs, we used logistic regression to evaluate the association between baseline abundance of each microbiome taxonomic group across phylogenies as well as pathways identified in our sequencing data. We identified substantial heterogeneity in our cohort in these microbial feature abundances, so we only analyzed pathways and taxa that occurred in 10 or more samples, yielding a total of 185 taxa and 2,213 pathways. Within these, we identified one statistically significant relationship after correcting for multiple hypothesis testing: decreased abundance of *Gemmiger formicilis* at endpoint and WBMs at endpoint (Beta coefficient = -1.02, q-value = 0.008). Otherwise, no taxa or pathway abundances were significantly associated with any clinical outcome variable after adjusting for False Discovery Rate (Supp Table 2, Fig 4).

**Figure 4:**
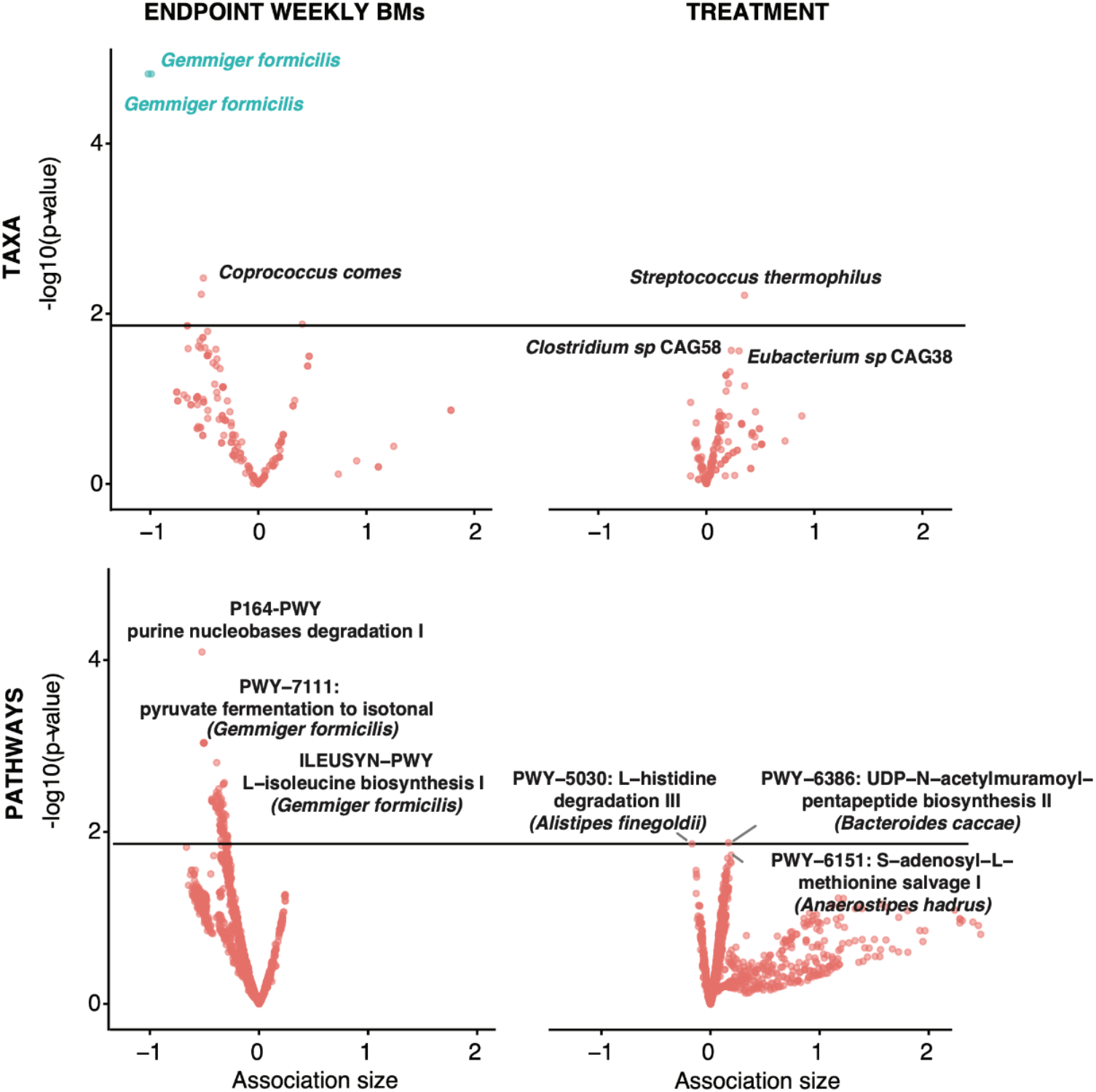
Associations with treatment and endpoint WBMs. X-axes are beta-coefficients from linear regressions. Y axes are negative log10(q-values). Color denotes FDR-significance, the solid line indicates a nominal p-value = 0.05. The 3 most significant associations are reported for each plot.

We additionally computed beta diversity between individuals in the treatment group at all phylogenetic levels (using MetaPhlAn3 output) and did not observe clear stratification by treatment status (Supp Fig 1).

### Response rate to probiotic treatment is contingent upon species richness

We next computed Shannon diversity. Simpson diversity, and taxonomic richness at the phylum, class, order, family, genus, and species levels for all samples, using Wilcoxon tests to compare variation in each of these three metrics between baseline and endpoint for non-responders and responders as well as individuals in the placebo group who both did or did not improve sans treatment Responders were defined as individuals who received treatment and experienced an increase in WBMs of ≥ 1. We did not see any association between response to treatment and Shannon or Simpson diversity.

However, we found that baseline richness of multiple phylogenetic levels (family, order, class, and phylum, p < 0.05 in all cases) discriminated between responders versus non responders (Fig 5). For these phylogenetic groups we additionally identified that richness increased between the baseline and endpoint timepoints (p < 0.05 in all cases, Supp Fig 2). Endpoint richness was not significantly different between responders and non-responders. We did not find that baseline richness could discriminate between individuals who received the placebo and improved on their own, nor did richness significantly change between baseline and endpoint for these individuals.

**Figure 5:**
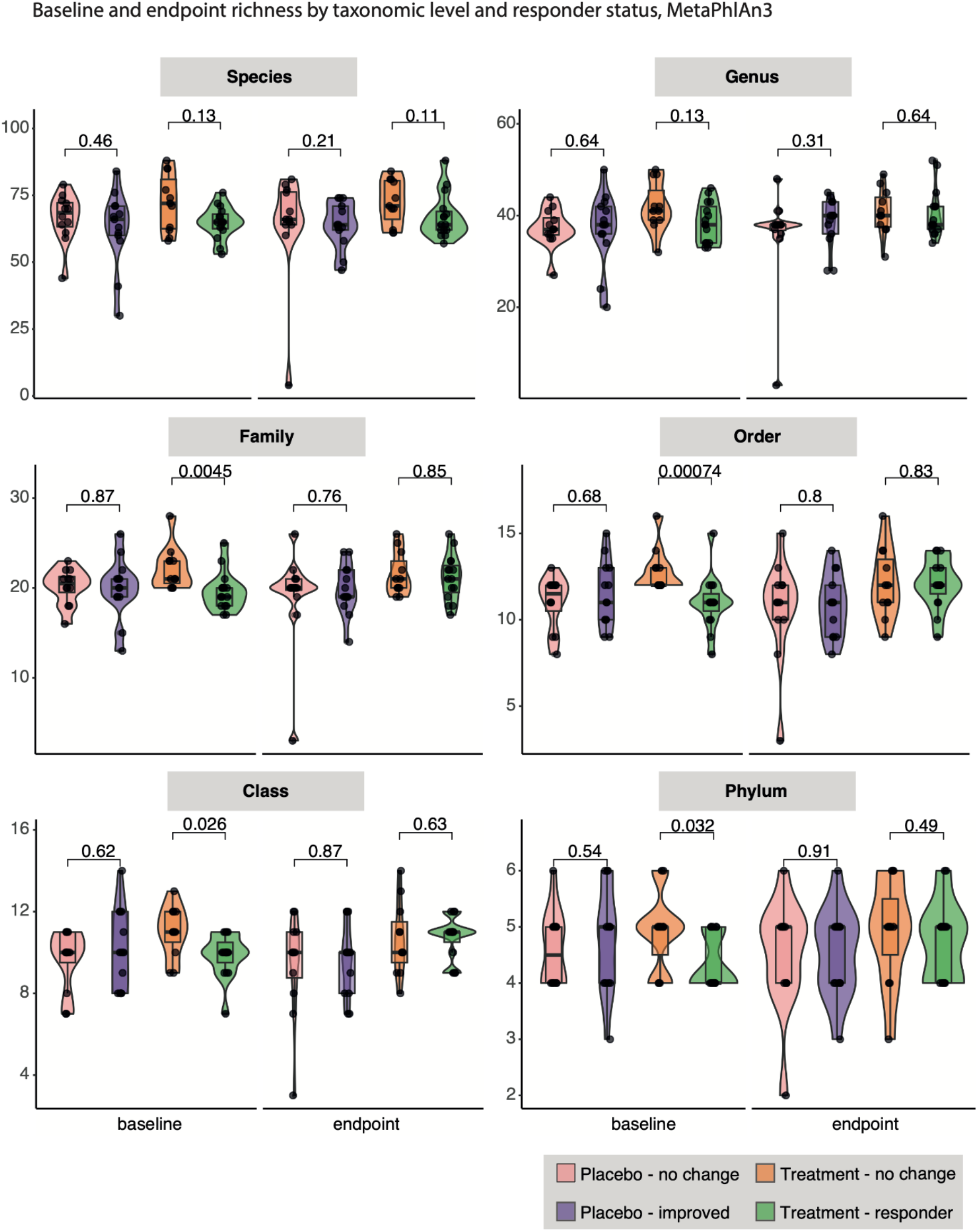
Response to treatment as a function of richness. We report how baseline richness changed as a function of future response to treatment. P-values derive from Wilcoxon tests.

We next stress-tested the ability of richness to discriminate between responders and non-responders using both alternative modeling approaches and a different method for quantifying taxonomic abundance within metagenomes (kraken2/bracken as opposed to MetaPhlAn3). We defined responders as individuals who received treatment and experienced an increase in WBMs of ≥ 1. Additionally, to account for potential confounding factors, we tested the association between richness at each phylogenetic level and responder status using a logistic regression approach adjusted for baseline WBMs and age. The association between responder status and richness was still either significant or trending towards significance for all groups (p < 0.05 or p < 0.1, Supp Table 3). The kraken2/bracken analysis yielded similar results (Supp Fig 3) as MetaPhlAn3, though different phylogenetic levels were statistically significant. We additionally used a logistic regression approach to evaluate the kraken2/bracken output and once more confirmed significance in association between richness and treatment response (Supp Table 3).

## Discussion

This placebo-controlled, randomized clinical trial demonstrated that a novel synbiotic formulation increased weekly WBMs in children who, at baseline, had low frequency WBMs. We additionally characterized the microbiome in individuals who received and responded to treatment versus those who did not, identifying microbial richness as an indicator of a high likelihood of response to treatment. Only a fraction of studies that evaluate the impact of microbial therapy on human health are executed in a placebo-controlled or randomized setting. While useful in many ways, non-randomized study designs are not able to test a fundamental causal link between treatment and disease. Moreover, high placebo response rates are typically observed in gastrointestinal trials with subjective endpoints due to the potential influence of stress, belief, and other psychosomatic influences on the gastrointestinal system and symptomology.^46^ In the case of adult IBS, for example, placebo response rates up to 40% are typically observed.^47^

Constipated children taking the synbiotic experienced a response rate comparable to trials testing the impact of fiber-based and laxative interventions on pediatric constipation. For example, a placebo-controlled recent study on polyethylene glycol (PEG) reported a response rate of up to 77% and a placebo response rate of 42%.^48^ These values are comparable to our reported treatment and placebo response rates (e.g. 62% and 13%, respectively, in the constipated, 2 WBM cohort). Our results were additionally similar to trials testing laxatives and other dietary interventions.^49,50^

Our cohort had two major drawbacks: (i) a number of non-constipated individuals not meeting our definition of constipation enrolled at baseline due to discrepancies between parental reporting of constipation and our clinical trial’s definition of constipation (being < 4 WBMs) and (ii) a high placebo response rate. Looking at the full cohort, we see a significant increase in the number of WBMs for both placebo and active. Having a statistically significant improvement in the placebo arm, generally referred to as a placebo effect, makes it more difficult to show superiority in the active arm. It is additionally worth nothing that placebos are difficult to design for microbiome clinical trials; for example, maltodextrin placebos (like the one used here) may have an impact on the gut microbiota, yielding a placebo response rate and raising the difficulty of observing a statistically significant response in the active group.^51,52^ Despite these limitations, however, we still observed a statistically significant response to treatment when considering the constipated cohorts, whereas non-constipated individuals taking the intervention experienced no significant change in WBMs as a function of treatment.

Additionally, one further limitation is that our analysis is based on a single nine-strain synbiotic composition in children. Patterns of microbial persistence upon use of other probiotic strains, or by populations not present in our study, such as infants, adults and individuals with preexisting medical conditions warrant further prospective human research. Also, bifidogenicity, strain persistence, and correlation with richness cannot be tied to individual strains, dosages, methods of delivery, or any other single feature of a microbial therapy.

This study expands our knowledge by employing several bioinformatics-based techniques to evaluate the effect of a rationally defined multi-species, multi-strain synbiotic in a pediatric population. We found no indication that the intervention adversely affected children and there were no reported adverse effects, increase in symptom severity, or dropout related to tolerability. Due to the impact of constipation on overall quality of life, it is encouraging that statistically significant improvements in ≥ 2 and ≥ 3 WBMs were found in constipated children, a response that we claim a medical practitioner would consider as clinically relevant.

We also were able to show that treatment affected individuals’ microbial gene composition. Specifically, in those who received treatment, we identified a potential bifidogenic effect as well as the persistence of all probiotic species across time. This was observed despite substantial heterogeneity in taxonomic signatures at baseline, and we did not see the same effect in the placebo group.

Additionally, despite limited individual microbial taxa being associated with clinical phenotypes writ large, we observed the ability of microbial richness to potentially predict -- across multiple phylogenetic levels, modeling approaches, and taxonomic characterization methods -- response to treatment. Shannon and Simpson diversity were not indicative of response. This yields implications for designing future trials related to personalized response based on an individual’s baseline microbiota. We hypothesize that the greater species richness is likely to translate directly into greater variation of functional traits and depletion of available resources, resulting in the competitive exclusion of exogenous bacteria. Other groups have identified personalized responses and resistance to microbial therapy, however the problem of identifying those individuals with a single indicator who are most likely to benefit remains open.^53^ We propose investigating richness as this indicator and hypothesize that the presence of potentially similar organisms or traits decreases the relative fitness and colonization opportunities of microbial therapy.

## Supporting information

Supplementary Table 1: Deidentified patient and sequencing metadata.

Supplementary Table 2: Results from taxonomic and pathway regressions.

Supplementary Table 3: Stress-testing the association between richness and responder status.

## Data Availability

All software used in this project is available at https://github.com/b-tierney/pds08. Due to the clinical nature of this study, data is available upon request.

## Supplement

**Supplementary Figure 1:**
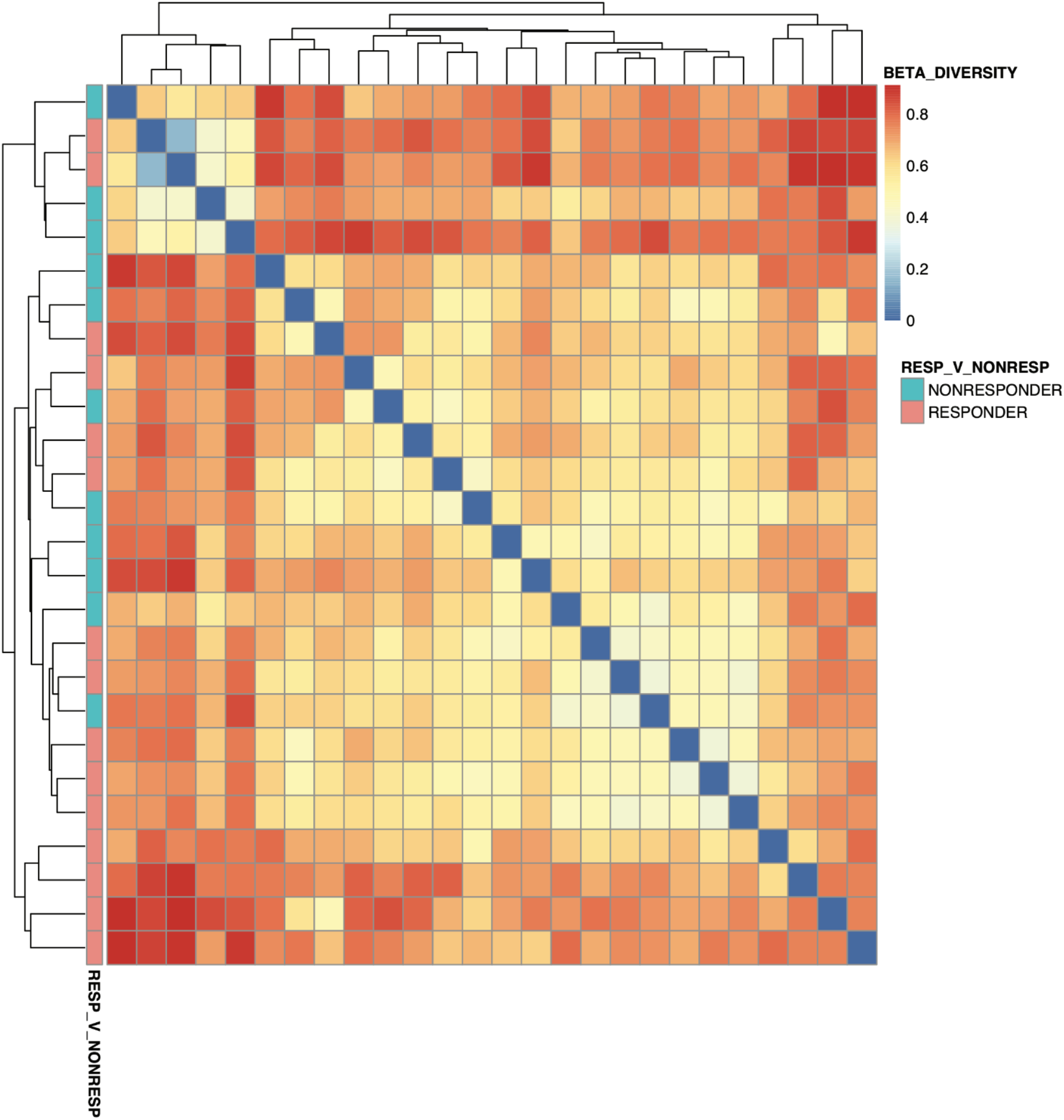
Beta diversity between responders and non-responders. Each row/column represents a different patient.

**Supplementary Figure 2:**
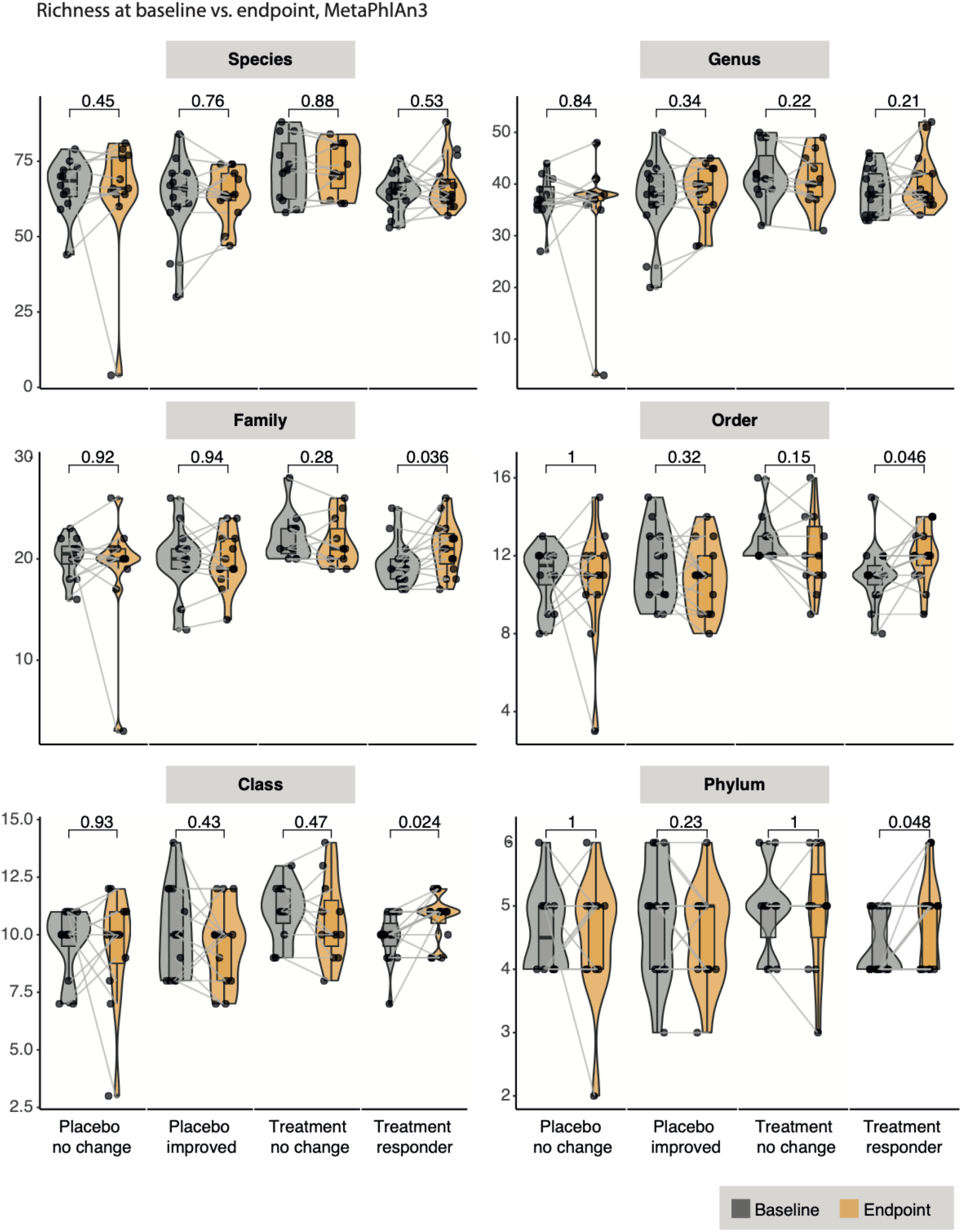
Change in richness between baseline and endpoints for all individuals across all taxonomic levels.

**Supplementary Figure 3:**
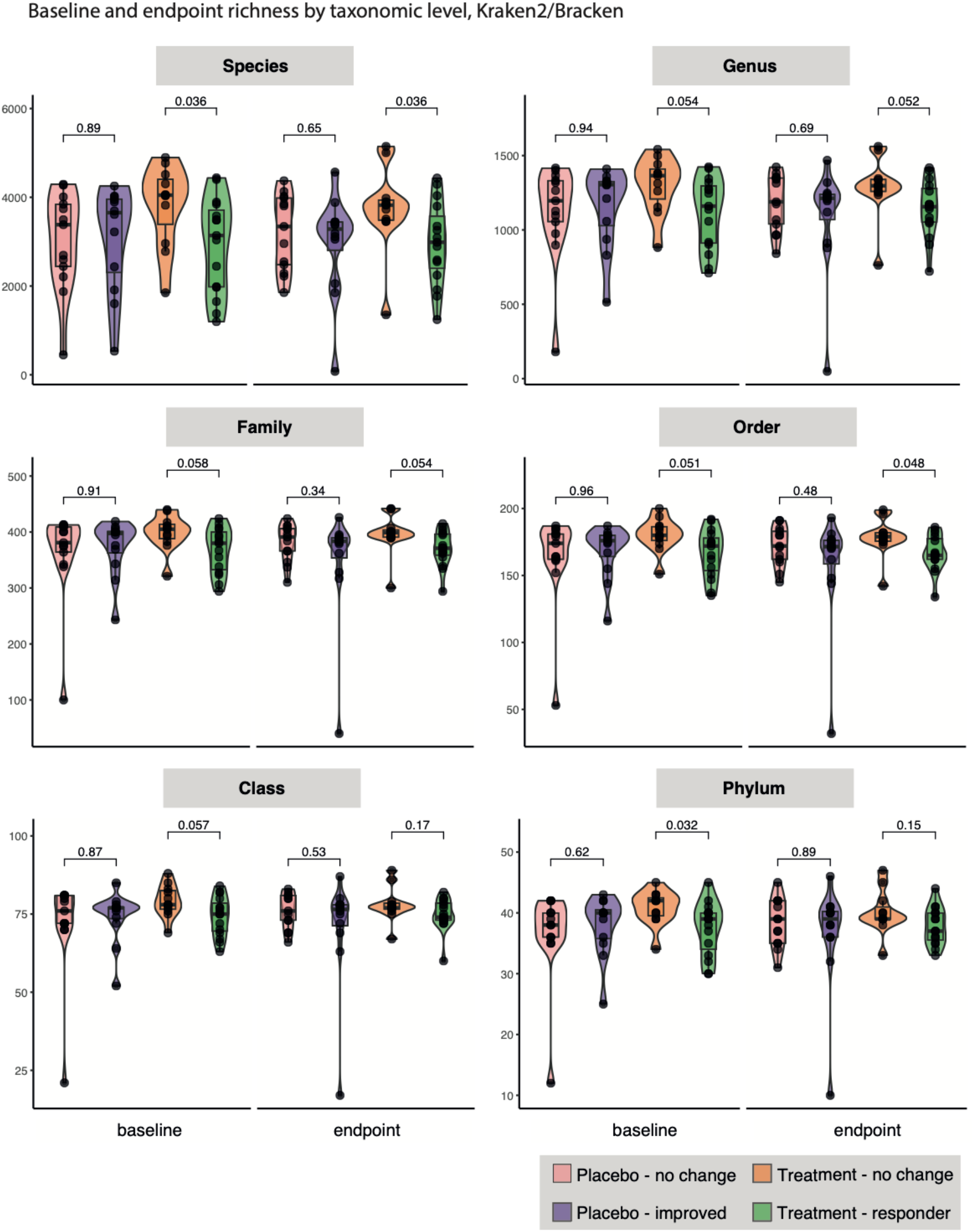
Comparing baseline richness across individuals using an alternative taxonomic classification approach, Kraken2/Bracken.

**Supplementary Table 1:** Deidentified patient and sequencing metadata.

**Supplementary Table 2:** Results from taxonomic and pathway regressions.

**Supplementary Table 3:** Stress-testing the association between richness and responder status. This table contains the output from logistic regressions for associating richness with response to synbiotic across different taxonomic association methods.

## Author contributions and acknowledgements

BTT led bioinformatic analysis and manuscript drafting without involvement in trial design. CEM and RD designed the bioinformatic strategy and participated in data analysis and manuscript drafting. BC contributed to manuscript review from a functional assessment and clinical perspective. JV contributed towards trial conception and text review. EC performed bioinformatics data analyses and visualization. GS assisted with statistical analysis. PAB contributed to data analysis and manuscript review from a prebiotic and probiotic perspective. SGP and NV contributed to data interpretation and manuscript review. JFP contributed to manuscript review with emphasis on bioinformatic analysis and computational techniques related to microbiome analysis. AF contributed towards manuscript review from a pediatric and clinical perspective. EAM contributed to manuscript review including pediatric and gastroenterological perspectives. JB provided clinical trial operations management and sample processing support. GR assisted with study design and manuscript writing.

## Competing interests

BTT and GS led data analysis and were not involved in study design or trial execution. JV, AF, EAM, CEM, and JFP are members of the Seed Health Scientific board. BTT, GS, NV, PAB, GR, SGP and EK are consultants for SH and were not involved in the microbial synbiotic or clinical trial design. BPC receives potential royalties from the Rome Foundation for usage of the modified Bristol Stool Form Scale for Children, which is not used in this study. AF is additionally a cofounder and stockholder of Alba Therapeutics, a company developing treatments complementary to the gluten-free diet by exploiting gut permeability. EAM is additionally on the Scientific Advisory Board of Axial Biotherapeutics, Pendulum Therapeutics, Bloom Science, Mahana Therapeutics, APC Ireland, Danone, and Amare. JFP is the Founder and Chief Scientific Officer of Diversigen, Inc. and is on the Scientific Advisory Board for 4D Pharma, PLC. CEM is co-founder of Biotia and Onegevity Health.

## Notes

### Clinical Trial

NCT04534036

### Funding Statement

This study was funded by Seed Health.

### Author Declarations

Ethics committee/IRB of IntegReview IRB gave ethical approval for this work.

